# Frontal Cortex-Subthalamic Nucleus Beta Oscillations Exhibit Phase Locking and Granger Causality in Parkinson’s Disease

**DOI:** 10.64898/2026.05.13.26348975

**Authors:** Jack Coursen, Toren Arginteanu, Gabriele Boccardo, Anruo Shen, Kelly A. Mills, Yousef Salimpour, William S. Anderson

## Abstract

**Objective:** Pathological beta oscillations are a hallmark of Parkinson’s Disease (PD) and are linked with symptom severity and therapeutic efficacy of deep brain stimulation (DBS). Although some studies suggest that beta oscillations may propagate from the frontal cortex to the subthalamic nucleus (STN), direct evidence based on cortical and subcortical neural recordings remains limited. This study investigates synchrony and directionality of beta-band interactions between the frontal cortex and STN in PD.

**Approach:** Simultaneous electrocorticography and STN local field potential recordings were obtained from three PD patients undergoing awake DBS lead placement surgery. Cortical-STN beta phase synchrony was quantified using phase locking value, and directed functional connectivity was analyzed using time-resolved bivariate Granger causality.

**Main results:** Phase locking value mapping revealed a spatially non-uniform distribution of beta phase synchrony, with the strongest coupling localized most prominently within the precentral and superior frontal gyri. Granger causality analysis demonstrated a predominance of cortical-to-subthalamic beta-band interactions across all subjects with intermittent bidirectional coupling.

**Significance:** These findings provide evidence that pathological beta oscillations in Parkinson’s may preferentially propagate from the frontal cortex to the basal ganglia, consistent with known motor pathways. These findings are consistent with a cortical contribution to pathological beta oscillations and highlight potential methods for obtaining cortical targets for phase-dependent neuromodulation.

## 1. Introduction

Parkinson’s disease (PD) is a neurodegenerative disease associated with loss of dopaminergic neuron function within the basal ganglia (BG) [1]. Resulting alterations in the cortico-BG-thalamo-cortical loop have been shown to cause abnormalities in narrow-band neural oscillations, such as excess beta band (13-30 Hz) power and synchronization [2], including excessive coupling between beta oscillation phase and gamma oscillation amplitude [3]. These signal abnormalities are often resolved by treatments such as deep brain stimulation (DBS) in association with a reduction of symptom severity [2][4][5]. Therefore, beta oscillation power and phase are of interest both for understanding the underlying pathology of PD and also as biomarkers to guide therapeutic interventions [4].

To better understand beta oscillations’ physiologic significance and therapeutic potential as biomarkers, prior work has sought to elucidate their cause and anatomical origin. According to one hypothesis, they emerge from network interactions between two nuclei of the BG: the subthalamic nucleus (STN) and globus pallidus externus (GPe) [1]. Alternatively, they may be caused by pathological changes in plasticity between neurons within the BG nuclei [1]. Delineating the direction of beta oscillation propagation through the cortical-BG circuitry could clarify their origin. They may originate in the cortex and propagate to the STN, thereby reaching the BG via the hyperdirect or indirect pathways [1]. Examination of signals by location within the STN has supported this hypothesis [6]. Further studies combining STN local field potentials (LFPs) with EEG and magnetoencephalography (MEG) data have revealed asymmetric beta interactions using spectral coherence and related measures, though with limited spatial and temporal specificity [7]. More recent invasive studies using spectral directed causality measures have identified interactions from the cortex to the STN at high-beta frequencies under dopaminergic therapy and DBS conditions [8].

We aim to better understand the relationship between beta oscillations in cortical and subcortical regions using simultaneous baseline electrophysiological recordings with high spatial and temporal resolution. We focus specifically on beta phase-based interactions between the frontal cortex and the STN, given their well-established yet incompletely understood role in abnormal PD oscillatory activity and their significance in phase-targeted neuromodulation therapies. We present a pilot study showcasing novel results leveraging a unique simultaneous cortical-subcortical recording and analysis.

## 2. Methods

### 2.1. Participants and Recording

Three PD patients undergoing awake bilateral STN DBS surgery were included. A 63-channel subdural electrocorticography (EcoG) electrode strip (PMT Corporation, Chanhassen, MN) was inserted over the sensorimotor cortex, while LFPs were simultaneously recorded from implanted microelectrodes targeting the STN (Alpha Omega, Nazareth, IS). ECoG signals were recorded digitally at a 1 kHz sampling frequency using a Neural Signal Processor (Blackrock Microsystems, Salt Lake City, UT), and STN LFPs were recorded at 1375 Hz using the Neuro Omega system (Alpha Omega, Nazareth, IS). Recordings were obtained during awake, resting-state conditions, and analyses were restricted to continuous segments of stable intraoperative recording (approximately 2 minutes per subject on average). Electrode locations were reconstructed using pre-implant MRI and intraoperative CT images, and were assigned neuroanatomical labels according to the Human Brainnetome Atlas using the FieldTrip and FreeSurfer toolboxes (Figure 1) [9][10].

**Figure 1.**
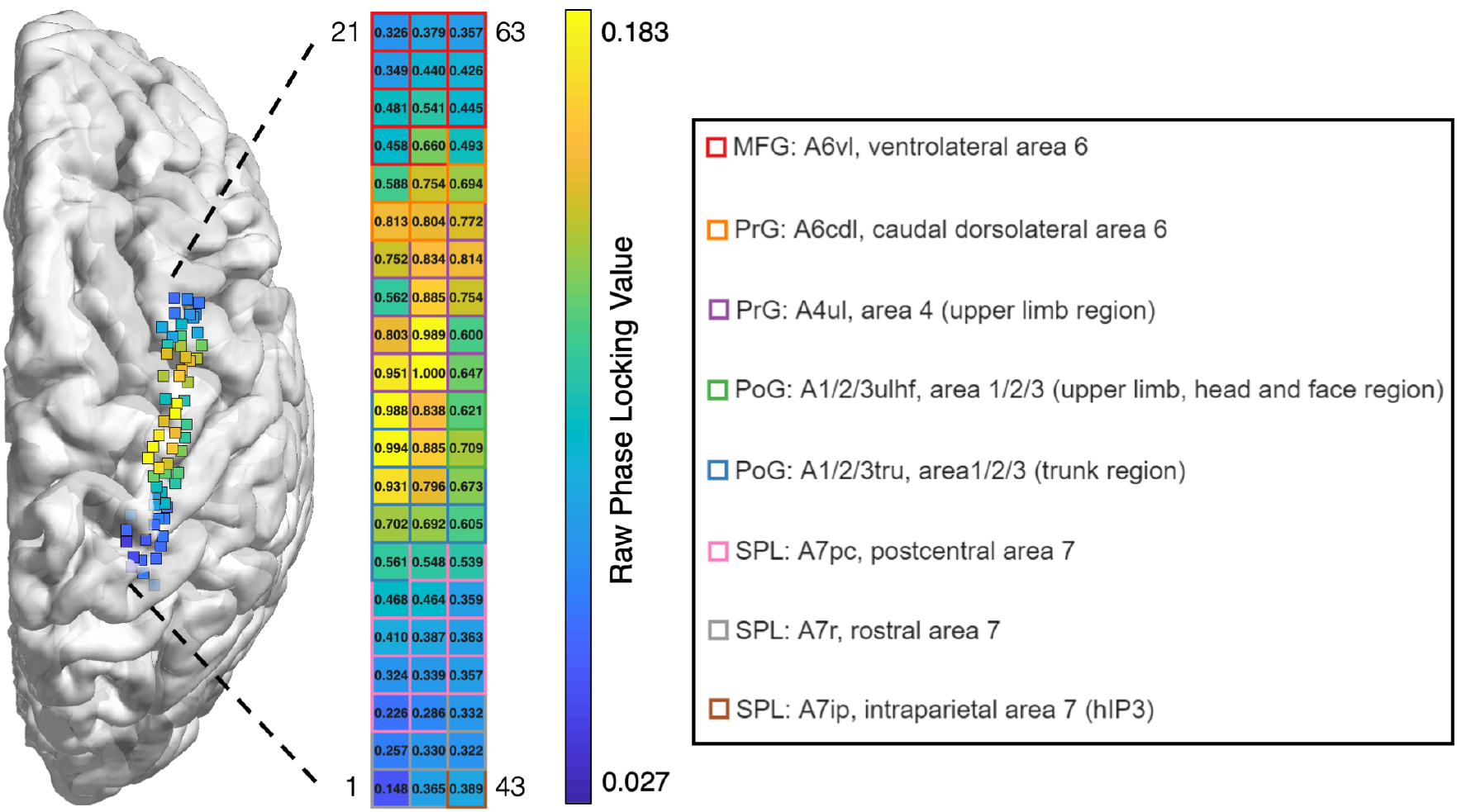
Visualization of Phase Locking Value (PLV) distribution for Subject 1. Left: Visualization of the ECoG array on the merged MRI and CT brain scans using the Montreal Neurological Institute (MNI) 3D coordinate system and FieldTrip MATLAB toolbox. Right: 2D heatmap of the PLV from STN to each cortical electrode channel with border color coding representing neuroanatomical labels according to the Human Brainnetome Atlas. *Abbreviations: MFG, middle frontal gyrus; PrG, precentral gyrus; PoG, postcentral gyrus; SPL, superior parietal lobule*.

### 2.2. Phase Locking Value

Phase Locking Value (PLV) quantifies the temporal phase variability between two signals over time, independent of signal amplitude, and ranges from 0 (no consistent phase relationship) to 1 (signals with a constant phase difference). PLV between two neural signals is computed using:

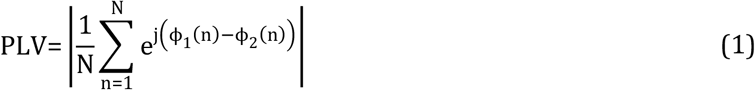

Where ϕ_1_(n) and ϕ_2_(n) are the phases of two signals at time n over N samples. Beta-band PLV between two channels were obtained by band-pass filtering signals to the beta frequency range (13-30 Hz), extracting instantaneous phase using the Hilbert Transform, and computing with (1). PLV was computed between all frontal cortex electrodes in relation to one STN LFP channel over continuous intraoperative recordings.

### 2.3. Granger Causality

Granger Causality (GC) is a measure of linear dependence that reflects directional predictability, testing whether the variance of error for a linear autoregressive (AR) model estimation of one signal can be reduced when using information from a second signal. If using knowledge of past information of the second signal (full model) improves the prediction of the first signal beyond information from the past values of the first signal alone (reduced model), there is said to be Granger causality from the second signal to the first. Larger GC values correspond to stronger functional connectivity, and a GC of 0 indicates no causality. Bivariate GC for two time-domain signals x and y can be computed using:

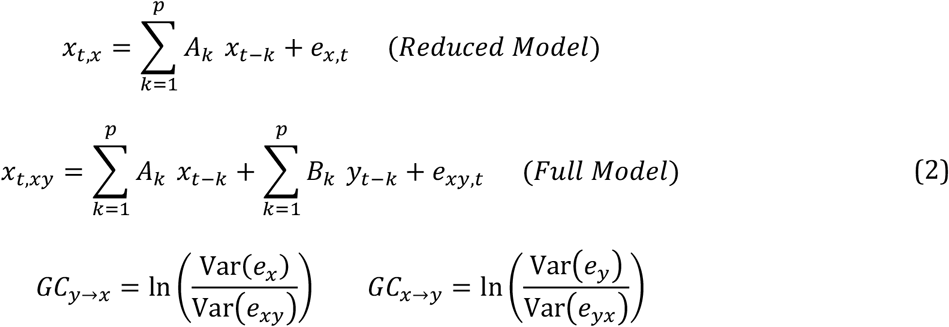

### 2.4. Statistical Analysis

All statistical analyses were performed in MATLAB (MathWorks, Natick, MA).

To assess whether observed PLV values exceed those expected by chance, results were compared with generated surrogate PLV data. Surrogate datasets were generated by circularly shifting the phase time series of the STN reference signal by a random temporal offset, while leaving the cortical phase time series unchanged. For each surrogate realization, PLV was recomputed between the ECoG channels with respect to the shifted STN phase using the previously discussed pipeline. This procedure was repeated for n = 100 independent realizations for each cortical-STN pair to provide an empirical null distribution for each electrode to help distinguish true phase locking from spurious connectivity. For each channel analyzed, one-sided empirical p-values were calculated from the proportion of realizations whose PLV was exceeded by the observed value, and statistical significance was assessed at α = 0.05. The proportion of electrodes exceeding their corresponding surrogate distribution means within anatomical regions was calculated descriptively.

Cortical electrodes with the highest PLV with respect to the STN signal were selected for GC analysis. GC was computed bidirectionally (cortex to STN and STN to cortex) in non-overlapping 250-millisecond windows (Figure 2). Within each subject, paired GC values from corresponding time windows (cortex-to-STN and STN-to-cortex) were compared using a two-tailed paired Wilcoxon signed-rank test (assessed at α = 0.05). Group-level summaries are presented with mean and standard error of mean (SEM) across the entire recording period for each subject.

**Figure 2.**
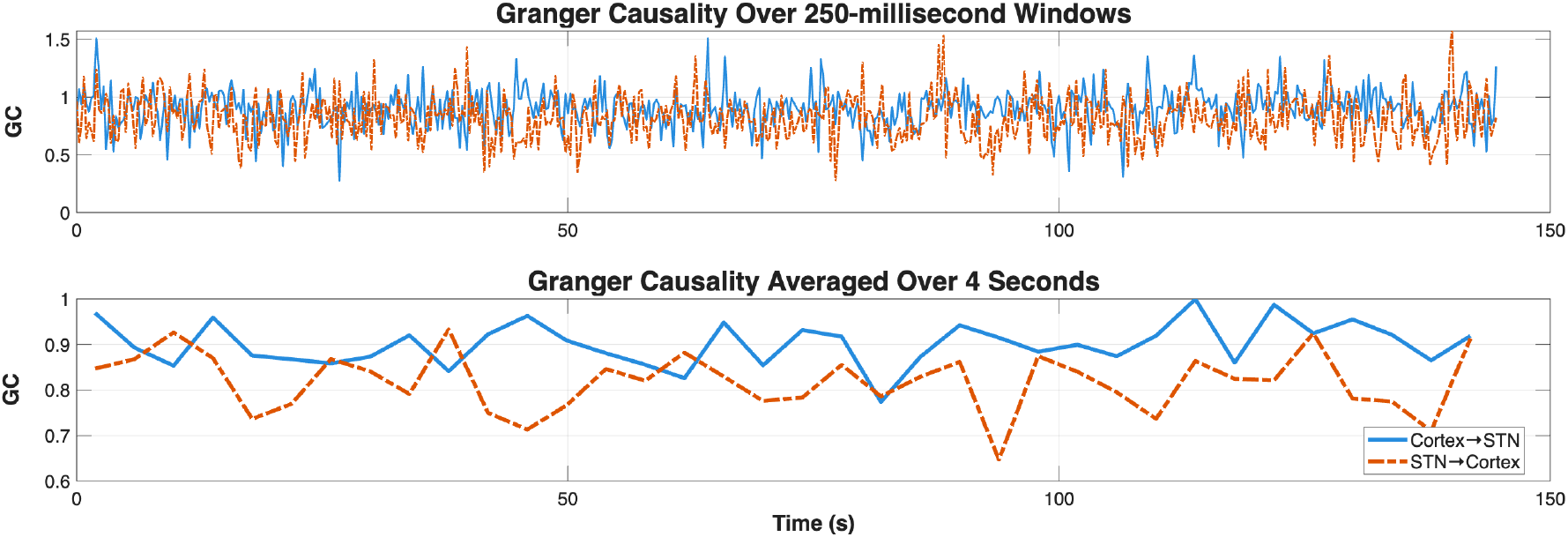
Time-resolved Cortical-Subthalamic Granger Causality (GC) Analysis for Subject 1. Top: (Cortical-to-Subthalamic (blue, solid) and Subthalamic-to-Cortical (orange, dashed) Granger Causality computed in consecutive, non-overlapping 250-millisecond windows over the entire intraoperative recording period. Bottom: Corresponding Granger Causality averaged over consecutive 4 second windows. *Abbreviations: GC, Granger Causality; STN, Subthalamic Nucleus*.

The Akaike Information Criterion (AIC) was used to select the model order with a ceiling of 50, corresponding to approximately one beta cycle at the central band frequency. The AIC was computed across GC windows to determine a suitable model order. One key assumption that enables GC analysis using linear AR models is that signals are stationary. To verify this assumption, the Augmented Dickey-Fuller (ADF) test was performed on the LFP and ECoG channel data used for analysis. The ADF test rejected the null hypothesis of a unit root for both signals (P < 0.001), indicating that these time windows can be approximated as stationary.

## 3. Results

Results were divided into analyses of cortical-subcortical beta phase locking and cortical-subcortical directed functional connectivity. To investigate whether beta oscillations exhibit preferential cortical–subcortical coupling and directed information flow, we first characterized the spatial distribution of beta phase synchrony between frontal cortical electrodes and STN LFPs. We then assessed the directionality of these interactions using time-resolved bivariate GC analysis.

### 3.1. Beta Phase Locking Analysis

Beta-band phase locking between the frontal cortex and STN was spatially non-uniform, with the highest PLV channels localized to the precentral and superior frontal gyri (Figure 1). Within these regions, elevated PLV values were distributed across multiple neighboring electrodes rather than being confined to a single focal site. Surrogate analysis demonstrated that the highest-PLV channels exhibited phase locking magnitudes that were statistically significant (Subject 1: *P* = 0.0099; Subject 2: *P* = 0.0099; Subject 3: *P* = 0.0495) relative to chance (Table 1). Additionally, 82.1% (23/28) of electrode channels within the precentral and superior frontal gyri exhibited PLV values greater than their corresponding surrogate averages. An example pairwise calculation of beta phase is presented in Figure 3.

**Table 1:** Spatial PLV and Surrogate Analysis Summary. The highest average Phase Locking Value (PLV) region’s value and label using the Human Brainnetome Atlas, the highest PLV cortical electrode channel value, and the PLV of its corresponding surrogate analysis distribution for all three patients is presented. Normalized PLV (NPLV) corresponds to phase locking values normalized to the highest PLV channel. *Abbreviations: PLV, Phase Locking Value; RPLV, Raw Phase Locking Value; NPLV, Normalized Phase Locking Value; ECoG, Electrocorticogram*.

| Subject | Highest Avg. PLV Region | Highest Avg. PLV Region |  | Highest PLV ECoG Channel | Highest PLV ECoG Channel Surrogate Distribution Avg. |  |
| --- | --- | --- | --- | --- | --- | --- |
|  |  | RPLV | NPLV |  | RPLV | NPLV |
| 1 | Right Precentral Gyrus | 0.1463 | 0.800 | 0.1829 | 0.0167 | 0.092 |
| 2 | Left Superior Frontal Gyrus | 0.0829 | 0.759 | 0.1092 | 0.0126 | 0.115 |
| 3 | Right Precentral Gyrus | 0.0249 | 0.530 | 0.0471 | 0.0202 | 0.429 |

**Figure 3.**
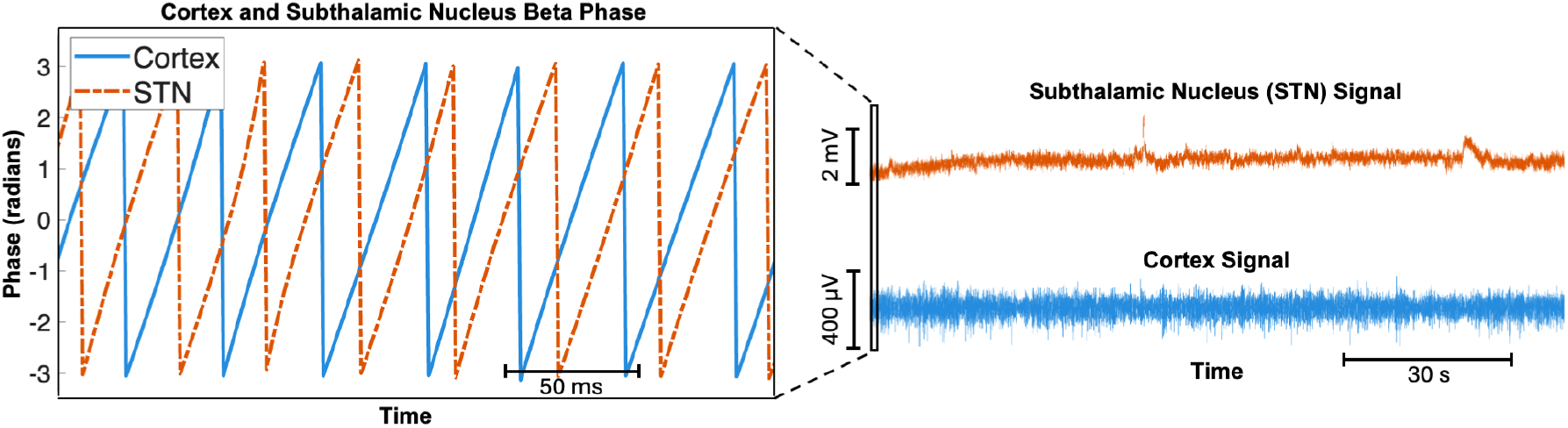
Beta-Xiltered signal and phase for Subject 1. Right: Beta-Xiltered STN LFP (orange, top) and ECoG (blue, bottom) signals across the intraoperative recording window. Left: An example 250-millisecond window of the corresponding phase time series, obtained using the Hilbert Transform of STN (orange, dashed) and ECoG (blue, solid) signals. Phase locking value for the highest cortical electrode in Subject 1 (channel 33) was 0.1829 during the example recording window.

### 3.2. Directed Functional Connectivity Analysis

Across consecutive, non-overlapping windows, time-resolved GC values from cortex-to-STN exceeded STN-to-cortex a majority of the time (Figure 2). GC in the cortex-to-STN direction was significantly greater than in the STN-to-cortex direction for all three subjects, as confirmed by paired within-subject Wilcoxon signed-rank testing (Subject 1: z = 7.565 *P* < 0.001; Subject 2: z = 10.377 *P* < 0.001; Subject 3: z = 9.326 *P* < 0.001). At a group level, mean GC values similarly demonstrated consistent predominance of cortical to subcortical functional connectivity (Table 2, Figure 4).

**Table 2:** Summary Granger Causality Statistics. The Cortex to STN and STN to Cortex Granger Causality mean and standard error of mean are presented for each subject. Corresponding z-statistics for the differences of these two directions according to paired Wilcoxon signed-rank testing results are presented. *Abbreviations: GC, Granger Causality; SEM, Standard error of the mean; STN, Subthalamic Nucleus*

| Subject | Cortex-STN GC |  | STN-Cortex GC |  | Wilcoxon Signed-Rank Test |
| --- | --- | --- | --- | --- | --- |
|  | Mean | SEM | Mean | SEM |  |
| 1 | 0.901 | 0.0068 | 0.8177 | 0.0082 | $z = 7.565^*$ |
| 2 | 0.8155 | 0.0058 | 0.7157 | 0.0088 | $z = 10.377^*$ |
| 3 | 1.0599 | 0.0105 | 0.9012 | 0.0109 | $z = 9.326^*$ |

\*Test statistics demonstrate statistical significance.

**Figure 4.**
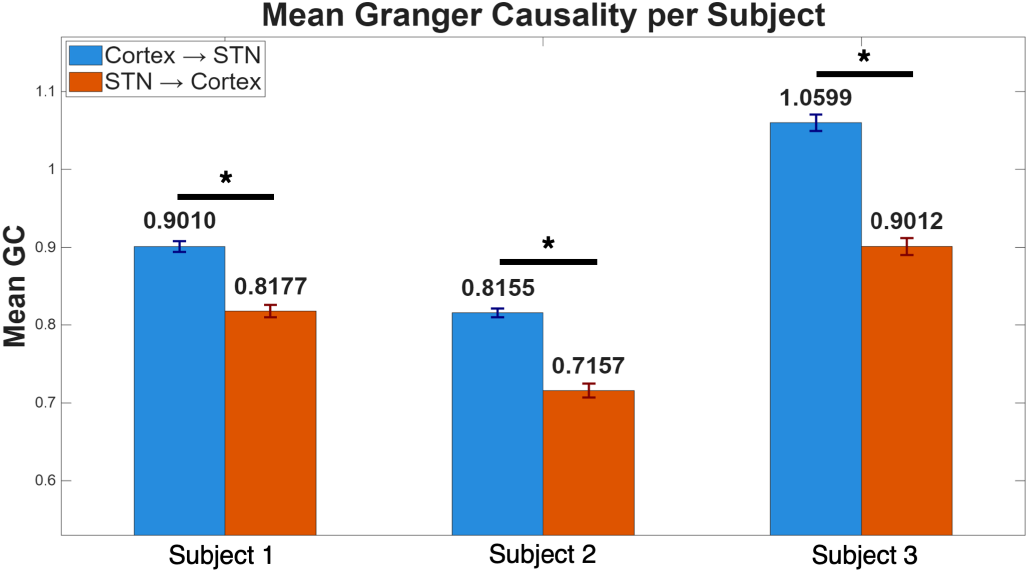
Summary of Granger Causality across all subjects. Cortical-Subcortical Granger Causality computed across all 3 three subjects with mean standard error bars. *Corresponding paired Wilcoxon signed-rank tests comparing Cortex-to-STN and STN-to-Cortex for each subject yielded statistically signiXicant differences[KM4.1] at an α = 0.05 signiXicance level (Subject 1: z = 7.565 P < 0.001; Subject 2: z = 10.377 P < 0.001; Subject 3: z = 9.326 P < 0.001). *Abbreviations: GC, Granger Causality; STN, Subthalamic Nucleus*.

## 4. Discussion

PLV mapping revealed that STN beta oscillations have unique synchronous relationships with different regions in the neocortex, and that such phase locking was significant relative to surrogates. The precentral gyrus and superior frontal gyri, regions within the primary motor and premotor cortex, most consistently demonstrated elevated beta synchrony with the STN. This finding is supported by existing histology, tractography, and electrophysiology studies, which describe connections from the primary and premotor cortex to the STN via the hyperdirect pathway [11].

GC analysis revealed that, for certain periods of time, cortical beta signals can help predict future values of STN signals, and to a lesser degree vice versa. This suggests that the direction of information flow between these two regions is bidirectional, with cortex-STN directed flow predominating over STN-cortex at rest. These results support the hypothesis that cortical pathological beta signals contribute substantially to STN beta dynamics within a recurrent cortico-basal ganglia network. The occasional reversal of GC was more sporadic and not statistically significant, and is consistent with a distributed, recurrent network with multiple pathways.

Although beta oscillation pathology is well established in PD, the network origin and degree of directionality of cortico-subthalamic interactions across patient states and frequency bands are unclear. Phase Locking Value [12] and Granger Causality have been used to study network interactions between electrophysiologic signals. Previous studies have debated whether pathological oscillations originate from within the basal ganglia, from within the cortex, or via network interactions between multiple regions [1]. In one study, both LFP and single neuron recording from within the STN were collected intraoperatively in PD patients [6]. This study determined that both predominant beta LFP power and the presence of cells firing in an oscillatory pattern within the beta frequency range were mainly found in the dorsal STN. Although this study did not record cortical signals, since the dorsal STN receives cortical input, these results may be consistent with a cortical drive hypothesis. Our findings affirm this, since we found that cortical beta signals appear to inform STN signals more than the converse, as measured by GC.

Furthermore, other studies have investigated information flow in simultaneous signals between the cortex and STN. Early studies combining STN LFPs with scalp EEG and MEG demonstrated asymmetric beta-band coupling using frequency-domain synchrony measures [7]. Later MEG-STN studies applied nonparametric spectral Granger causality to demonstrate predominant cortical-to-STN beta band directionality during brief rest windows, and STN-to-cortex directionality emerging in the gamma band during movement [13]. More recently, larger-scale invasive studies have strengthened evidence for directed cortico-STN beta interactions in PD by exploring spectral GC measures under medication and stimulation conditions [8]. Notably, it was found that STN deep brain stimulation selectively suppressed cortical drive to the STN in the high beta band without increasing that in the reverse direction. This supports a model in which the amplitude of pathological beta oscillations flowing from the cortex is disrupted under stimulation.

Our study builds upon these results by instead looking at spatially resolved cortical synchronization and time-resolved directionality over extended baseline intraoperative windows. Whereas the previous studies examined GC based on the broadband amplitude spectrum, our time domain analysis captures both phase and amplitude characteristics of the signal to examine the temporal persistence of beta asymmetries while focusing on the limited band of interest. Our results affirm the previous studies’ findings of notable information flow from cortex to STN, especially those which examined functional connectivity during resting conditions [13], and incorporation of resolved spatial phase synchrony adds an additional dimension to analyses relevant for localizing common or patient-specific regions of interest. Beta-gamma phase amplitude coupling is a specific biomarker of PD symptom severity [14]; therefore, taking beta phase into account offers additional vital information on the propagation of a pathological signal. Therefore, our spatial and temporal analyses may provide more insight into the unique spatial organization of pathological interactions and better predict the efficacy of neuromodulation techniques that require time-resolved stimulation protocols and personalized localization of stimulation targets.

Thus, this work is significant in that it elucidates the direction of propagation of a well-known pathological signal and lends evidence to support one hypothesis that has been debated in the literature. Our findings can help clarify the phase-based network interactions that produce this phenomenon, which in the future may lead to a better understanding of how it precisely relates to the symptoms of PD. Based on our results, motor pathways such as the hyperdirect pathway (or the indirect pathway through other intermediaries) may play a prominent role in the propagation of pathological beta oscillations from the cortex to the STN. Diffusion-weighted MRI tractography studies have demonstrated structural connectivity between motor cortical regions and the STN and investigated its use for predicting clinical outcomes following DBS [15]. Future work combining patient-specific tractography with our study’s synchrony and directed connectivity measures may help confirm whether structural connectivity relates to the magnitude and directionality of such beta-band interactions.

Beyond mechanistic insight, this work provides guidance on potential targets for therapeutic brain stimulation. Given its potential as a biomarker of PD symptom severity, the abnormal beta oscillation has been examined as a target for closed-loop neuromodulation approaches [4]. In particular, phase-targeted stimulation presents a promising avenue to directly modulate the pathological coupling of beta oscillations in PD when signal phases are known [14][16]. PLV mapping provides a means of identifying patient-specific cortical regions that are strongly phase-locked with the STN, which could serve as candidate sites for cortical sensing or stimulation. These regions could be used for estimating phase dynamics in deep brain structures, which may inform or guide closed-loop phase-dependent stimulation with the intent of modulating cortical-subcortical phase dynamics less invasively. The information flow observed using GC from cortex to STN also affirms the potential efficacy of cortical stimulation approaches aimed at influencing BG activity.

Studies have already shown that cortical stimulation targeting specific oscillation phases can impact disease-relevant physiology biomarkers [13]. Future investigation should determine whether inhibitory cortical stimulation can disrupt the transmission of pathological beta oscillations to the STN, and whether such stimulation could have therapeutic benefits comparable to STN-DBS. If successful, this could provide a much less invasive alternative therapy. Future work may build upon this pilot study by analyzing simultaneous ECoG and STN recordings in a larger cohort. Granger causality offers a powerful data-driven framework for identifying directional dependencies; however, work exploring direct mechanistic causation through lesioning studies and animal models of PD can provide additional validation of inferred network dynamics. Additionally, extending analyses to include single-cell spiking activity may help bridge observed oscillatory phenomena with their underlying cellular network.

## 5. Conclusion

This pilot study provides evidence that pathological beta oscillations in Parkinson’s Disease exhibit bidirectional information flow with preferential propagation from the frontal cortex to the subthalamic nucleus. Cortical regions within the precentral and superior frontal gyri exhibited strong beta phase synchrony with STN local field potentials, while Granger Causality analysis demonstrated predominant cortical-to-subthalamic information flow. These findings support an origin of beta oscillations consistent with known motor pathways and highlight potential for use of phase locking and Granger Causality to guide patient-specific phase-dependent neuromodulation therapies.

## Acknowledgements

The authors express gratitude towards the participants, surgical team, and Johns Hopkins Neuromodulation and Advanced Therapies Center. The authors do not have any conflicts of interest to disclose.

## Ethics Statement

This study was performed in accordance with the Declaration of Helsinki. This human study was approved by the Johns Hopkins Medicine Institutional Review Board. All adult participants provided written informed consent to participate in this study.

## Data Availability

The data that support the findings of this study are available from the corresponding author upon reasonable request.

